# Motivation, intention and action: wearing masks to prevent the spread of COVID-19

**DOI:** 10.1101/2022.05.25.22275599

**Authors:** Geoff Kaine, Vic Wright, Suzie Greenhalgh

**Affiliations:** Manaaaki Whenua Landcare Research, Hamilton, New Zealand; University of New England, Armidale, New South Wales, Australia; Manaaaki Whenua Landcare Research, Auckland, New Zealand

## Abstract

Governments around the world are seeking to slow the spread of COVID-19 by implementing measures that encourage, or mandate, changes in people’s behaviour. These changes include the wearing of face masks, social distancing, and testing and self-isolating when unwell. The success of these measures depends on (1) the willingness of individuals to change their behaviour and (2) their commitment and capacity to translate that intention into actions. Consequently, understanding and predicting the willingness of individuals to change their behaviour, and their enthusiasm to act on that willingness, is critical in assessing the likely effectiveness of these measures in slowing the spread of the virus.

In this paper we analyse responses to two separate regional surveys about people’s intentions and behaviour with respect to preventing the spread of COVID-19 in New Zealand. While motivations and intentions were largely similar across the regions, there was marked difference in action across the regions, specifically with respect to the frequency of wearing face masks. Our analysis suggests that the translation of intention (preventing the spread of COVID-19) into action (as measured by self-reported frequency of face mask use) was strongly associated with perceptions of the risk of infection (as measured by regional case numbers).

The results highlight the importance to policy design of distinguishing the factors that might influence the formation of behavioural intentions from those that might influence the implementation of those intentions.

## Introduction

The success of measures to slow or stop the spread of COVID-19 such as wearing face masks and social distancing, depends on the commitment and capacity of individuals to comply with them, and change their behaviour accordingly [1, 2, 3]. Ineffective compliance with these measures can put the achievement of policy outcomes at risk [4, 5]. For example, failure to wear face masks and socially distance may put the outcome of slowing the spread of COVID-19 at risk and may mean considerable resources must be invested in enforcement to avoid increased rates of infection, higher mortality, and the imposition of lockdowns causing both economic and psychological damage.

Understanding and predicting the extent to which individuals are willing to change their behaviour is critical, then, in assessing how effective measures like mask wearing and social distancing are likely to be, and whether alternatives such as curfews and lockdowns can be avoided. Unfortunately, good intentions do not necessarily translate into action. Hence, understanding why individuals may not change their behaviour, despite their good intentions, is also crucial if policies to encourage measures like wearing face masks and social distancing are to be effective.

Building on previous research [6, 7, 8, 9], we draw on the social psychological concepts of involvement and attitudes in this paper to predict the willingness of the public in several regions across New Zealand to prevent the spread of COVID-19 by wearing face masks, self-isolating when unwell, and getting tested for COVID-19. We then investigate differences across the regions in the extent to which intentions to wear face masks translated into mask wearing behaviour. Our aim was to explain why, despite similarity in people’s intentions with respect to preventing the spread of COVID-19, there were dramatic differences in their behaviour with respect to wearing face masks.

## Theory

Research and analysis focused on the process underlying decisions that direct the actions of individuals has long included recognition that there are two phases to the process. The natural point of separation between these phases is the ‘action intention’ which arises once the decision is made. This action intention is normally referred to as ‘behavioural intention’ in the literature [10, 11, 12 13]. Behavioural intention, rather than actual behaviour, is usually the appropriate outcome on which to focus when seeking to understand decision-making because actual behaviour can diverge from intended behaviour for a wide variety of reasons. Seeking to relate the actual behaviour to decision processes triggered by a decision problem would unhelpfully conflate decision considerations and implementation considerations, masking the contribution of the former. In effect, the contemporary approach is to distinguish decision making from decision implementation.

While factors that may impact upon implementation could well figure in the decision-making process (particularly as costs or benefits attaching to decision options), substantive modelling of implementation is required to complete the analysis, or projection, of actual behaviour flowing from behavioural choices made in response to perceived decision problems.

In any specific applied setting, particularly those involving existing practices and products, decision implementation is routine and familiar to all users. In the case of new practices and products, decision implementation assumes greater importance because it defines the rate of adoption of the novel item. Measures such as wearing face masks and social distancing that were introduced to contain the spread of COVID-19 fit into this novel category: when introduced these practices were new to each person, even the advice to wash hands much more regularly is novel, and the wearing of face masks obviously so. Hence, when new practices are being introduced to cope with a pandemic, whose impacts are driven by rates of transmission across a population, rates of practice adoption matter a good deal (as reflected by concerns about ‘vaccine hesitancy’ [14, 15]).

In prior studies [6, 7, 8, 9] we investigated behavioural intentions, and actual behaviour, with respect to measures advocated by government to suppress or eliminate COVID-19 in New Zealand. These analyses using the I_3_ model [16] assigned people to segments according to their involvement with specific government policy outcomes (eliminating COVID-19 from New Zealand) and related policy measures (wearing face masks, self-isolating, getting tested for COVID-19, using a COVID-19 tracer app, and getting vaccinated for COVID-19).

The segments in each study were analysed to identify, within each segment, the diversity and roles of salient beliefs and attitudes, and whether segment members were favourably or unfavourably predisposed to comply with the policy measures. This approach was designed to identify policy and promotional activity that could be implemented to strengthen desired behavioural intentions with respect to the policy measures (in the context of current behavioural intentions, and salient attitudes and beliefs). The research was not designed to model decision-making processes per se.

In two studies [6, 7] in different regions, behavioural intentions with respect to wearing face masks, self-isolating and getting tested for COVID-19 were investigated together with self-reports of actual behaviour with respect to wearing face masks and getting tested. The results in these two studies indicated that, despite similarities across regions in behavioural intentions, there were marked dissimilarities in actual behaviour [7]. In particular, although the willingness to act to prevent the spread of COVID-19 was similar across the regions, the wearing of face masks was dramatically different across the regions [7].

Where, as in this instance, diversity in actual behaviour occurs in a context of shared behavioural intentions with respect to a novel behaviour, it is necessary to identify the cause. First, because, if the claimed integrity of the I_3_ model [6, 7, 9, 16, 17, 18, 19, 20], and other models of behavioural intentions [10, 11], is to be sustained, it is necessary to be able to identify a plausible and active cause for hesitancy to implement an intention. In a basic sense, behavioural intention is not the policy target if it is detached in some major way from actual behaviour. Second, because, for any additional actions to be taken to accelerate adoption of the behaviour to be appropriate, the causes of the hesitancy need to be identified.

Bagozzi [10], one of the few theorists to model the implementation of behavioural intentions, has suggested there is a planning and control loop that can be identified which governs the translation of intention into (ongoing) behaviour. This loop determines the ‘how’ and ‘when’ of implementation, and the monitoring of initial actions regarding the adequacy of goal attainment and any unanticipated outcomes. This outcome information is fed back into the decision-making and implementing system.

Bagozzi [10] draws attention to the fact that different sets of, sometimes partly overlapping, factors influence the formation of behavioural intentions and their implementation. This has consequences for understanding what factors can properly be said to act as ‘barriers’ [10] to desired behaviour changes. The notion of a ‘barrier’ is often unhelpfully broad because, as here, the set of factors serving to create a favourable or unfavourable attitude to novel behaviour can, and in this case must, be different to the set of factors seeming to impede behaviour change. Particularly, ‘barriers’, in normal usage, usually refers to things or situations impeding movement in an intended direction: intended by the subject, not some observer. ‘Barriers’ has relevant meaning, therefore, to factors causing behavioural intention to not lead to the behaviour in question. Its use with respect to the forming of behavioural intentions reveals more about observer preferences than impediments the subject confronts.

In the case of mask wearing, the barrier to continuous action may be non-availability of masks, unanticipated social opprobrium when they are worn or unexpected discomfort (both of which reflect poor judgement in arriving at behavioural intention). The most obvious, and logically the first, ‘barrier’ to seek out is the absence of a behavioural trigger. Identifying behavioural intention by questioning subjects rather than tracking behaviour, is to discover a predisposition to act. A failure to act implies the absence of a trigger to activate the predisposition. In this case, if masks are readily available, socially acceptable, and reasonably comfortable, the missing trigger will presumably be related to perceived need: the perceived threat of airborne infection.

The perceived threat of airborne infection is, inevitably, subjective and cue-driven amongst most people. The cues employed to judge the threat of infection may well be influenced by reported infections in an area, and trends in them, and perhaps by the prevalence of mask wearing. Infection incidence may be the critical catalyst that triggers action. The adoption of behaviours such as the wearing of face masks has been associated with a range of variables including perceptions of the perceived risk of infection, the local incidence rate of COVID-19 and feelings of stress in relation to COVID-19 [21].

In the next section we provide a brief description of the history of COVID-19 in New Zealand to place the subsequent analysis in its proper context.

## COVID-19 in New Zealand

COVID-19 was first detected in New Zealand on 28 February 2020 [22]. Within three weeks the central government had closed New Zealand’s international border to all except returning citizens and permanent residents. The government began pursuing a restrictive strategy [23] of eliminating COVID-19 and applied a range of controls (policy measures) to stop the transmission of COVID-19 in New Zealand [24]. Elimination (the desired policy outcome) did not necessarily mean eradicating the virus permanently from New Zealand; rather, that central government was confident chains of transmission in the community had been eliminated for at least 28 days, and any cases imported from overseas in the future could be effectively contained [24].

The central government instituted a four-tier alert system that mandated policy measures such as: progressively tighter restrictions on people’s movement outside their homes and immediate families, including travelling to work; social distancing and encouraging the wearing of masks outside the home at the higher alert levels; and self-isolating and seeking testing if people felt unwell or experienced symptoms characteristic of COVID-19 infection [22].

On 25 March 2020, New Zealand moved to a Level 4 ‘lockdown’, the highest level of alert, and a National State of Emergency was declared [22]. At this alert level people are instructed to stay at home except for essential personal movement such as for health care or essential shopping, safe recreational activity is allowed in the local area, and travel is severely limited. All gatherings are cancelled, and all public venues closed. Businesses are closed except for essential services (for example, supermarkets, pharmacies, health clinics, petrol stations and lifeline organisations). All educational facilities are closed [22].

As the spread of the virus slowed and stopped, the country progressively moved to lower alert levels: Level 3 towards the end of April and Level 2 in early May 2020. Alert Level 1, the lowest level, was introduced on 8 June 2020 because community transmission had halted and there were no active cases in the country outside the Managed Isolation and Quarantine facilities (MIQ). These were established specifically to quarantine all incoming travellers to New Zealand for 14 days after arrival. If a traveller tested positive for COVID-19 at any time during the 14 days, they were moved to another quarantine facility for people with COVID-19 [22].

However, on 11 August 2020 four new cases were detected in Auckland. Auckland returned the next day to Alert Level 3, with the rest of the country at Alert Level 2. Auckland remained at Alert Level 3 until 30 August, when it moved to Level 2, with additional restrictions on travel and the size of gatherings. The rest of the country remained at the standard Alert Level 2 until 21 September, when the alert level was downgraded to Level 1. The extra restrictions on Auckland residents were relaxed on 21 September and they returned to Alert Level 1 on 7 October 2020 [24].

The central government commenced a mass vaccination programme for COVID-19 using the Pfizer vaccine, starting with border staff and managed isolation and quarantine workers, in February 2021 [9, 25]. The programme was accompanied by an extensive, government-funded publicity campaign using traditional and social media.

## Materials and methods

Data from two surveys were used in this study. The first survey, the ‘Auckland’ survey, was of Auckland residents and was conducted over two weeks from 7 September to 22 September 2020. Auckland was chosen for the survey because it is New Zealand’s largest city and is the mostly likely place for community transmission to occur, given the greater number of MIQ facilities and frontline border workers in the city. At the time of the survey, Auckland residents were mostly under Alert Level 2, which meant that they were expected to maintain social distancing when outside their homes and to wear masks in public places. They were also expected to keep track of their movements and to self-isolate and seek testing for COVID-19 if they felt unwell and experienced symptoms associated with COVID-19.

The second survey, the ‘regional’ survey, was of residents in five regions outside Auckland with, or near, MIQ facilities (Hamilton, Rotorua, Tauranga, Wellington, and Christchurch) and was conducted during the first and second week of March 2021, before the Delta variant was detected in New Zealand and before vaccinations were available to the general public. When the survey commenced, residents in these regions were under Alert Level 2, which meant that they were expected to maintain social distancing when outside their homes and to wear masks in public places. They were also expected to keep track of their movements and to self-isolate and seek testing for COVID-19 if they felt unwell and experienced symptoms associated with COVID-19. On 8 March 2021 the regions shifted to Alert Level 1, which meant people were expected to wear masks on public transport. They were also encouraged to keep track of their movements and to self-isolate and seek testing for COVID-19 if they felt unwell and experienced symptoms associated with COVID-19 [22].

For both surveys, a questionnaire seeking information from the public on their beliefs about, attitudes towards, and willingness to wear face masks, self-isolate and be tested for COVID-19 was designed based on the I_3_ Compliance Framework [16]. Involvement was measured using a condensed version of the Laurent and Kapferer [26] involvement scale developed by Kaine [27], with respondents rating two statements on each of the five components of involvement (functional, experiential, identity-based, risk-based, and consequence-based).

Attitudes were measured using a simple, evaluative scale (the questionnaire is reproduced in S1). The ordering of the statements in the involvement and attitude scales was randomised to avoid bias in responses. Respondents indicated their agreement with statements in all the involvement, attitude and belief scales using a five-point rating, ranging from strongly disagree (1) to strongly agree (5).

Respondents’ propensity to wear face masks was obtained by asking them if they had worn a face mask when out in public the previous week and whether they had to go out to work the previous week. Respondents answered both questions using a five-point scale ranging from ‘always’ to ‘never’. Their propensity to self-isolate was obtained by asking them, ‘Thinking about the next few days, would you stay home if you were unwell or had any of the following symptoms: a dry cough, fever, loss of sense of smell, loss of sense of taste, shortness of breath or difficulty breathing?’. We also asked, ‘If you were advised to do so by a healthcare professional or public health authority, would you self-isolate for 14 days?’. Both questions were answered using a five-point scale ranging from ‘definitely’ to ‘definitely not’.

Information was also sought on the demographic characteristics of respondents, including age, education, and ethnicity, and whether they wore masks, would self-isolate and had been tested for COVID-19. The ethnicity categories were Māori (the Indigenous people of New Zealand), European New Zealander, Pacific Islander, Asian and Other.

Participation in surveys was voluntary, respondents could leave the survey at any time, and all survey questions were optional and could be skipped. The research approach was reviewed and approved by the Manaaki Whenua – Landcare Research’s social ethics process (application no. 2021/10 NK) which is based on the New Zealand Association of Social Science Research code of ethics.

The Auckland questionnaire had been piloted with a small random sample of residents (n=30), and subsequently completed by a larger random sample of residents (n=1001) who were members of a large-scale, commercial consumer internet panel. The regional questionnaire, which was identical to the Auckland questionnaire, was completed by a large random sample of residents (n=2000), stratified by regional population, who were also members of a large-scale, commercial consumer internet panel. Panel members receive reward points (which are redeemable for products and services) for completing surveys. For both surveys, an internet link to the questionnaire was distributed to randomly selected members of the panel subject to the constraint that they were resident in the relevant region and were not minors. To reduce respondent fatigue, a split-sample approach was taken, with each respondent answering sets of questions in relation to two of the three measures (mask wearing, self-isolation and getting tested for COVID-19).

Given the pandemic had been receiving widespread coverage by mainstream and social media in New Zealand since February 2020, it seems reasonable to suppose that virtually all the residents in each region were aware of COVID-19 at the time of the surveys and that most, if not all, were also aware of the government claims of the social desirability of wearing face masks and social distancing when out in public, and of self-isolating if feeling unwell and getting tested for COVID-19.

While awareness of the existence of COVID-19 is a prerequisite for involvement with it, awareness does not necessarily entail involvement. Widespread awareness of COVID-19 simply creates the potential for widespread involvement. The extent to which that potential is realised depends on respondents’ beliefs about how COVID-19 could affect the achievement of their functional, experiential and self-identity needs.

Involvement scores were computed for each respondent as the simple arithmetic average of their agreement ratings for the 10 statements in the involvement scales. Attitudes scores were computed as the simple arithmetic average of their agreement ratings for the five statements in the attitude scales. For this paper, data was also gathered on the number, dates, and location by District Health Board of COVID-19 cases reported by the New Zealand Ministry of Health [28].

Statistical analyses were conducted using SPSS [29].

## Results

As the data on involvement, attitudes, behavioural intentions, and behaviour were collected using the same survey but at different times, a number of confounding factors may give rise to dissimilar results. These are:

• differences in the demographic composition of the samples
• differences in Alert Level restrictions
• differences in awareness of the prevalence, or nature, of COVID-19.

To begin with, although the samples were broadly similar with regard to their age, education, and income composition, they differed substantially with respect to gender and ethnicity (see Tables 1, 2, 3 and 4). Approximately 53% of respondents to the Auckland survey were women whereas approximately 65% of respondents to the regional survey were women.

**Table 1.** Age distribution of respondents

| Age category (years) | Percentage of Auckland respondents | Percentage of regional respondents |
| --- | --- | --- |
| 18–29 | 22.8 | 13.2 |
| 30–39 | 21.8 | 22.6 |
| 40–49 | 18.4 | 21.5 |
| 50–59 | 13.1 | 12.5 |
| 60–69 | 12.5 | 13.8 |
| 70 and over | 11.4 | 16.4 |

**Table 2.** Distribution of respondents by highest educational qualification.

| Education category | Percentage of Auckland respondents | Percentage of regional respondents |
| --- | --- | --- |
| Some or all of secondary school | 14.2 | 19.5 |
| Certificate (1–6) | 12.4 | 19.1 |
| Diploma (5–7) | 14.3 | 17.5 |
| Bachelor | 33.6 | 23.4 |
| Post-graduate diploma/certificate | 10.2 | 11.1 |
| Post-graduate degree | 15.3 | 9.4 |

**Table 3.**
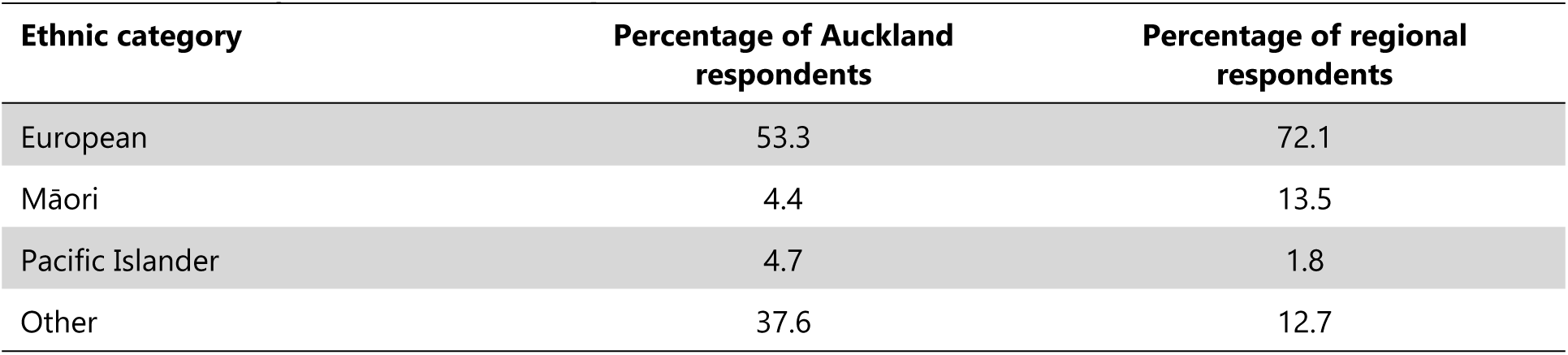
Ethnicity distribution of respondents.

| Ethnic category | Percentage of Auckland respondents | Percentage of regional respondents |
| --- | --- | --- |
| European | 53.3 | 72.1 |
| Māori | 4.4 | 13.5 |
| Pacific Islander | 4.7 | 1.8 |
| Other | 37.6 | 12.7 |

**Table 4.** Income distribution of respondents.

| Income category | Percentage of Auckland respondents | Percentage of regional respondents |
| --- | --- | --- |
| Less than \$20,000 | 4.3 | 8.5 |
| \$20,000 to \$50,000 | 21.2 | 26.0 |
| \$50,000 to \$70,000 | 18.6 | 21.7 |
| \$70,000 to \$100,000 | 22.0 | 22.1 |
| More than \$100,000 | 33.8 | 21.6 |

There were statistically significant but weak associations [30] between the demographic characteristics and willingness to take responsibility for eliminating COVID-19 and willingness to change normal behaviour, make sacrifices and work with others to eliminate COVID-19 (see Table 5). There were also some statistically significant but weak associations [30] between demographic characteristics and the wearing of face masks and willingness to self-isolate (see Table 6). Differences in demographic characteristics such as age, ethnicity, gender, and education may influence perceptions of the danger to health posed by COVID-19. This suggests that differences in the demographic composition of the two surveys could partly explain differences in intentions and behaviour in the two surveys.

**Table 5.** Demographics and behavioural intentions.

| Characteristic | Feel some responsibility | Change normal behaviour | Work with others | Make sacrifices |
| --- | --- | --- | --- | --- |
| Age | 0.006* | 0.007* | 0.019* | 0.014* |
| Gender | 0.003* | 0.005* | 0.003* | 0.005 |
| Education | 0.009* | 0.006* | 0.008* | 0.006* |
| Ethnicity | 0.006* | 0.003 | 0.003 | 0.001 |
| Income | 0.016* | 0.014* | 0.009* | 0.011* |

Values are $\eta^2$ , the proportion of the variance in intention explained by the variance in the socio-demographic variable [30]. For example, differences in income explain 1.4% of the variation in intention to change normal behaviour ( $\eta^2 = 0.014$ ).
\* Indicates statistically significant effect ( $p < 0.01$ ).

**Table 6.** Demographics, wearing face masks and intention to self-isolate.

| Characteristic | Wore a face mask in public | Wore a face mask at work | Stay home if unwell | Stay home if instructed by health authority |
| --- | --- | --- | --- | --- |
| Age | 0.011* | 0.016* | 0.039* | 0.029* |
| Gender | 0.009* | 0.016* | 0.007* | 0.004 |
| Education | 0.021* | 0.029* | 0.004 | 0.003 |
| Ethnicity | 0.052* | 0.066* | 0.009* | 0.015* |
| Income | 0.002 | 0.005 | 0.010* | 0.014* |

Values are $\eta^2$ [30]

We investigated the effect of differences in Alert Level restrictions by classifying regional respondents into three categories according to the date when they completed the questionnaire. The first category consisted of respondents who completed the questionnaire prior to 8 March 2021, the date at which Alert Levels changed (from 2 to 1). The second category consisted of respondents who completed the questionnaire between 8 March 2021 and 15 March 2021, who could have been reporting on their behaviour under either Alert Level 1 or 2 depending on how they interpreted the phrase ‘Did you wear a face mask whenever you went out in public last week?’. The third and final category consisted of respondents who completed the questionnaire after 15 March 2021, and so should have been reporting on their behaviour under Alert Level 1.

We found a statistically significant, but extremely small, difference between the categories in the mean scores for willingness to change normal behaviour (ƞ2=0.007), make sacrifices (ƞ2=0.006) and work with others to eliminate COVID-19 (ƞ2=0.007). We also found a statistically significant, but extremely small, difference between the categories in the mean scores for wearing face masks in public (ƞ2=0.012) and wearing face masks at work (ƞ2=0.11). This suggests that the difference in Alert Levels between the two surveys may also partly explain differences in intentions and behaviour in the two surveys.

It seems unlikely that differences between the surveys in intentions and behaviour could be the result of differences in awareness of the prevalence, or nature, of COVID-19. Almost 730 cases of COVID-19 were detected in Auckland prior to the Auckland survey [28]. Approximately 570 cases had been detected in the MIQ regions prior to conducting the regional survey [28]. As noted earlier, the pandemic received extensive media coverage, including daily daily government briefings, in New Zealand during the six months preceding the Auckland survey. The Delta variant of COVID-19, a more highly infectious and severe variant of the virus [31, 32], emerged during the five months between the Auckland and regional surveys but was not present in New Zealand at the time the regional survey was conducted. Given the emergence of the Delta variant, and that the regional survey occurred some five months after the Auckland survey, awareness of COVID-19 among regional respondents could reasonably be expected to be at least as great as it was among Auckland respondents.

## Intentions and behaviour

The purpose of this analysis was to explain differences in the propensity of Auckland and regional respondents to comply with wearing face masks in public and at work, given their behavioural intentions were similar. Note that satisfactory reliabilities [33] were obtained in both surveys for the involvement and attitudinal scales with respect to eliminating COVID-19, wearing face masks, self-isolating when unwell and getting tested for COVID-19 (see Table 7).

**Table 7.** Reliability of involvement and attitude scales.

|  | Auckland | Regional |
| --- | --- | --- |
| Involvement with eliminating COVID-19 | 0.847 | 0.853 |
| Involvement with wearing face masks | 0.852 | 0.863 |
| Attitude towards wearing face masks | 0.854 | 0.814 |
| Involvement with self-isolating when unwell | 0.707 | 0.758 |
| Attitude towards self-isolating when unwell | 0.795 | 0.800 |
| Involvement with getting tested for COVID-19 | 0.821 | 0.807 |
| Attitude towards getting tested for COVID-19 | 0.849 | 0.813 |

Notes: Values are Cronbach's alpha [33].

The means for all the involvement, attitude, intention, and behaviour variables for both surveys are reported in Table 8. Where the means for the two surveys were statistically significantly different, the magnitude of the differences, as measured by effect size [30], is also reported in the table. An inspection of the table reveals that wearing face masks in public and at work were the only variables for which the means were statistically significantly different, and the magnitude of the difference was large, for the two surveys.

**Table 8.**
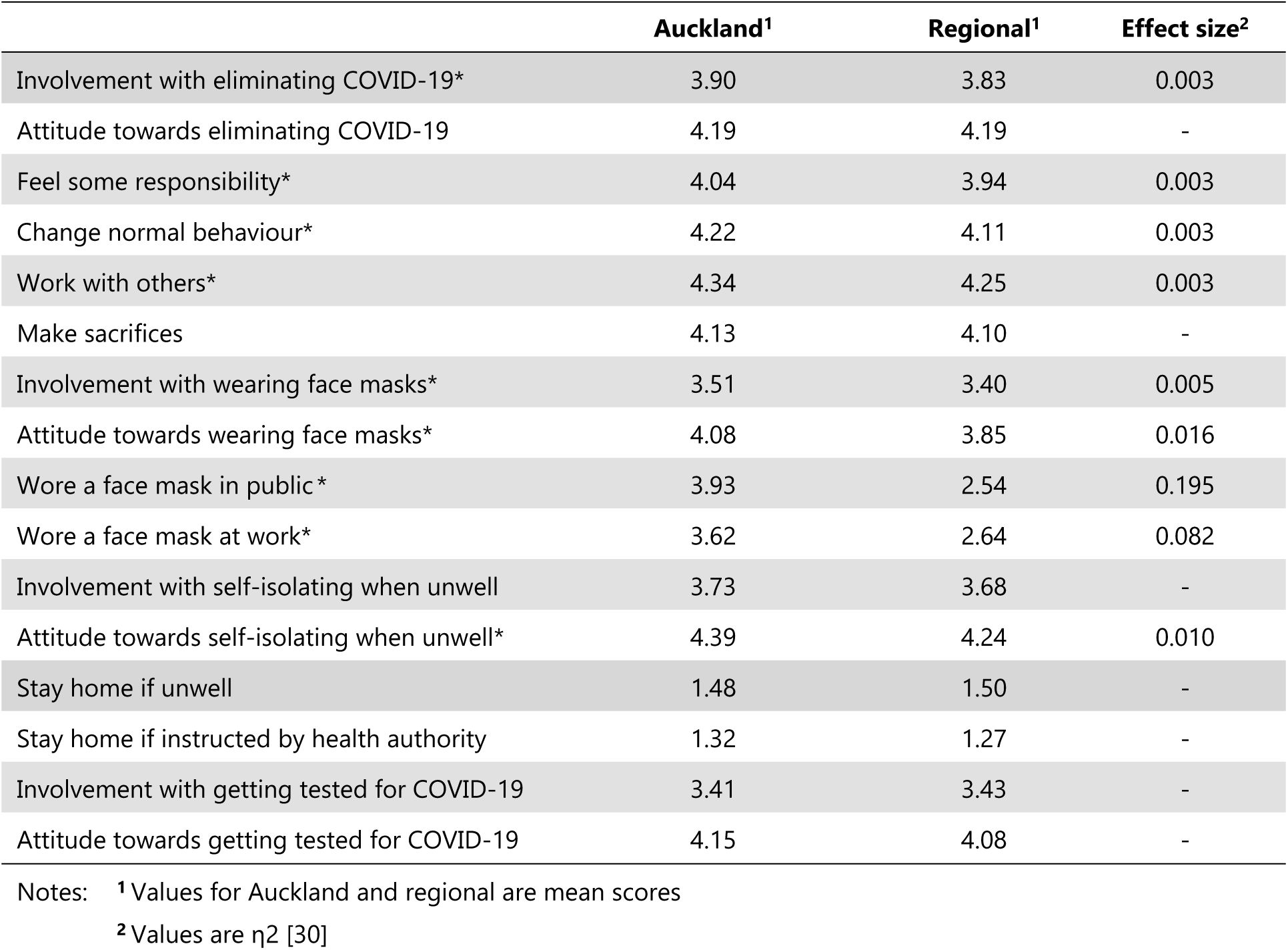
Involvement, attitude, intentions, and behaviour.

These results suggest that, on average, respondents in both surveys were similar in their motivations (as measured by involvement) and attitudes towards eliminating COVID-19, wearing face masks, self-isolating and getting tested for COVID-19. They were similar, on average, regarding their intentions to take some responsibility for eliminating COVID-19, and their intentions to change their normal behaviour, work with others and make sacrifices to eliminate COVID-19 from New Zealand. They were also similar, on average, regarding their intentions to self-isolate and get tested for COVID-19. Relatedly, Auckland and regional respondents were similar in their patterns of beliefs about COVID-19, eliminating COVID-19 from New Zealand, and the advantages and disadvantages of wearing face masks, self-isolating and getting tested for COVID-19 [7].

The only substantive difference between the two samples relates to the wearing of face masks, with the means for the regional sample being substantially lower than the means for the Auckland sample. The difference, on average, roughly corresponds to regional respondents reporting that they only wore face masks sometimes (at best) when they were out in public whereas Auckland respondents reported they often wore face masks when they were out in public.

## Predicting intentions and behaviour

Following Kaine et al. [6, 7] we hypothesised that respondents’ intentions with respect to eliminating COVID-19 from New Zealand would be a function of their involvement with, and attitude towards, eliminating COVID-19. Consequently, we estimated regressions with respondents’ willingness to take some responsibility for eliminating COVID-19, and their willingness to change normal behaviour, work with others and make sacrifices to eliminate COVID-19 from New Zealand as dependent variables.

The explanatory variables were respondents’ involvement with, and attitude towards, eliminating COVID-19. To account for the possibility that the demographic differences between the surveys might be correlated with relevant omitted variables (e.g. social norms, susceptibility to infection, risk of severe symptoms), respondents’ demographic characteristics were also included in the regressions. Dummy variables were created representing respondents’ ethnicity with ‘Other’ treated as the benchmark. Two additional dummy variables were included in the regressions, one representing the period when regional respondents were at Alert Level 2 and one representing the first week regional respondents were at Alert Level 1.

The explanatory power of the regressions, and the resulting parameter estimates, are reported in Table 9. The regressions were statistically significant and, for cross-sectional data, a substantial proportion of the variance in respondents’ intentions was explained by their involvement with, and attitude towards, eliminating COVID-19. The results show that involvement and attitude account for the bulk of the explained variation in the dependent variables. The variation in respondents’ intentions were only weakly related, if at all, to their demographic characteristics or, for regional respondents, changes in Alert Levels.

**Table 9.** Standardised parameter estimates for behavioural intentions.

|  | Feel responsible for<br>eliminating COVID-19<br>(n=2586) | Prepared to change<br>normal behaviour<br>(n=2586) | Willing to make<br>sacrifices<br>(n=2586) | Willing to work<br>together<br>(n=2586) |
| --- | --- | --- | --- | --- |
| **Involvement with eliminating COVID-19** | 0.466<br>($p<0.001$) | 0.479<br>($p<0.001$) | 0.399<br>($p<0.001$) | 0.469<br>($p<0.001$) |
| **Attitude towards eliminating COVID-19** | 0.232<br>($p<0.001$) | 0.272<br>($p<0.001$) | 0.353<br>($p<0.001$) | 0.295<br>($p<0.001$) |
| **Age** | 0.044<br>($p=0.007$) | 0.044<br>($p=0.005$) | 0.062<br>($p<0.001$) | 0.058<br>($p<0.001$) |
| **Gender** | 0.037<br>($p=0.017$) | 0.047<br>($p=0.001$) | 0.023<br>($p=0.118$) | 0.052<br>($p<0.001$) |
| **Education** | 0.040<br>($p=0.011$) | -0.008<br>($p=0.593$) | -0.015<br>($p=0.333$) | 0.002<br>($p=0.916$) |
| **Income** | 0.081<br>($p<0.001$) | 0.034<br>($p=0.027$) | 0.004<br>($p=0.817$) | 0.045<br>($p=0.003$) |
| **European NZ** | -0.049<br>($p=0.009$) | -0.020<br>($p=0.272$) | -0.018<br>($p=0.315$) | 0.000<br>($p=0.984$) |
| **Māori** | -0.051<br>($p=0.004$) | -0.042<br>($p=0.014$) | -0.053<br>($p=0.002$) | -0.023<br>($p=0.166$) |
| **Pacific Islander** | -0.004<br>($p=0.801$) | -0.009<br>($p=0.566$) | -0.016<br>($p=0.281$) | 0.019<br>($p=0.187$) |
| **Regions at Alert Level 2** | -0.013<br>($p=0.386$) | -0.040<br>($p=0.007$) | -0.002<br>($p=0.875$) | 0.001<br>($p=0.938$) |
| **First week regions at Alert Level 1** | -0.012<br>($p=0.453$) | 0.000<br>($p=0.996$) | -0.003<br>($p=0.842$) | 0.015<br>($p=0.320$) |
| **Adjusted R<sup>2</sup>** | 0.43 | 0.48 | 0.48 | 0.50 |
| **F-Test significance** | <0.001 | <0.001 | <0.001 | <0.01 |

Again, following Kaine et al. [6, 7], we hypothesised that respondents’ propensity to self-isolate and wear face masks would be a function of their involvement with, and attitude towards, self-isolating and wearing face masks, respectively. Consequently, we estimated regressions with respondents’ self-reported willingness to self-isolate and wearing of face masks as dependent variables. The explanatory variables for willingness to self-isolate were respondents’ involvement with, and attitude towards, wearing face masks and self-isolating.

Again, to account for the possibility that the demographic differences between the surveys might be correlated with relevant omitted variables, respondents’ demographic characteristics were also included in the regressions as well as dummy variables representing when regional respondents were at Alert Level 2 and the first week at Alert Level 1.

Willingness to self-isolate when unwell, or instructed to do so by a health authority, was strongly and positively influenced by involvement with, and attitudes towards, self-isolating (see Table 10). As with the other behavioural intention variables, the variation in respondents’ intentions to self-isolate were only weakly related, if at all, to their demographic characteristics or, for regional respondents, changes in Alert Levels.

**Table 10.** Standardised parameter estimates for intention to self-isolate.

|  | **Intending to stay home if**<br>**feeling unwell**<br>**(n=1697)** | **Stay home if instructed**<br>**to do so**<br>**(n=1697)** |
| --- | --- | --- |
| **Involvement with self-isolating** | 0.131<br>(*p*<0.001) | 0.100<br>(*p*<0.001) |
| **Attitude towards self-isolating** | 0.330<br>(*p*<0.001) | 0.399<br>(*p*<0.001) |
| **Age** | 0.174<br>(*p*<0.001) | 0.090<br>(*p*<0.001) |
| **Gender** | 0.086<br>(*p*<0.001) | 0.041<br>(*p*=0.072) |
| **Education** | 0.038<br>(*p*=0.103) | 0.001<br>(*p*=0.958) |
| **Income** | 0.043<br>(*p*=0.058) | 0.036<br>(*p*=0.115) |
| **European NZ** | 0.038<br>(*p*=0.159) | 0.016<br>(*p*=0.556) |
| **Māori** | 0.009<br>(*p*=0.718) | -0.032<br>(*p*=0.214) |
| **Pacific Islander** | 0.032<br>(*p*=0.158) | -0.016<br>(*p*=0.486) |
| **Regions at Alert Level 2** | -0.036<br>(*p*=0.112) | -0.055<br>(*p*=0.014) |
| **First week regions at Alert Level 1** | -0.029<br>(*p*=0.209) | 0.017<br>(*p*=0.441) |
| **Adjusted R<sup>2</sup>** | 0.21 | 0.23 |
| **F-Test significance** | <0.001 | <0.001 |

The explanatory variables for wearing face masks were respondents’ involvement with, and attitude towards, wearing face masks and, as with previous regressions, respondents’ demographic characteristics were also included to account for the possibility that the demographic differences might be correlated with relevant omitted variables. As before, we also included dummy variables representing when regional respondents were at Alert Level 2 and the first week at Alert Level 1.

We included additional explanatory variables in the regressions for wearing masks which were intended to account for the differences observed previously in the wearing of face masks by Auckland and regional respondents. We had attributed this difference in behaviour to differences in respondents’ perceptions of the risk of COVID-19 infection. We hypothesised that respondents would, in some way, use information about the number of cases in their region in forming their perception of the risk of being infected with COVID-19.

To begin with, we assumed that respondents’ perception of the risk of infection would be proportional to the total number of COVID-19 cases reported in their region prior to the survey, and that their perception of the risk of infection would become increasingly sensitive to the total number of cases, as that total increased. Consequently, the two additional variables included in the regressions were the total number of cases in a respondent’s region, and the total number of cases in their region squared (and centred to avoid multi-collinearity). Note that respondents might also use Alert level settings in judging the risk of infection.

We then assumed that respondents’ perception of the risk of infection would be proportional to the total number of COVID-19 cases reported in their region prior to the survey expressed as a fraction of the population of the region, and that their perception of the risk of infection would become increasingly sensitive to the number of cases expressed as a fraction of the population of the region, as the total number of cases increased. Consequently, the two additional variables included in these regressions were the total number of cases in a respondent’s region expressed as a proportion of the regional population, and the total number of cases expressed as a proportion of the regional population squared (and centred to avoid multi-collinearity). Noting again that respondents might also use Alert level settings in judging the risk of infection, the results, with and without the additional risk perception variables, are reported in Tables 11 and 12.

**Table 11.**
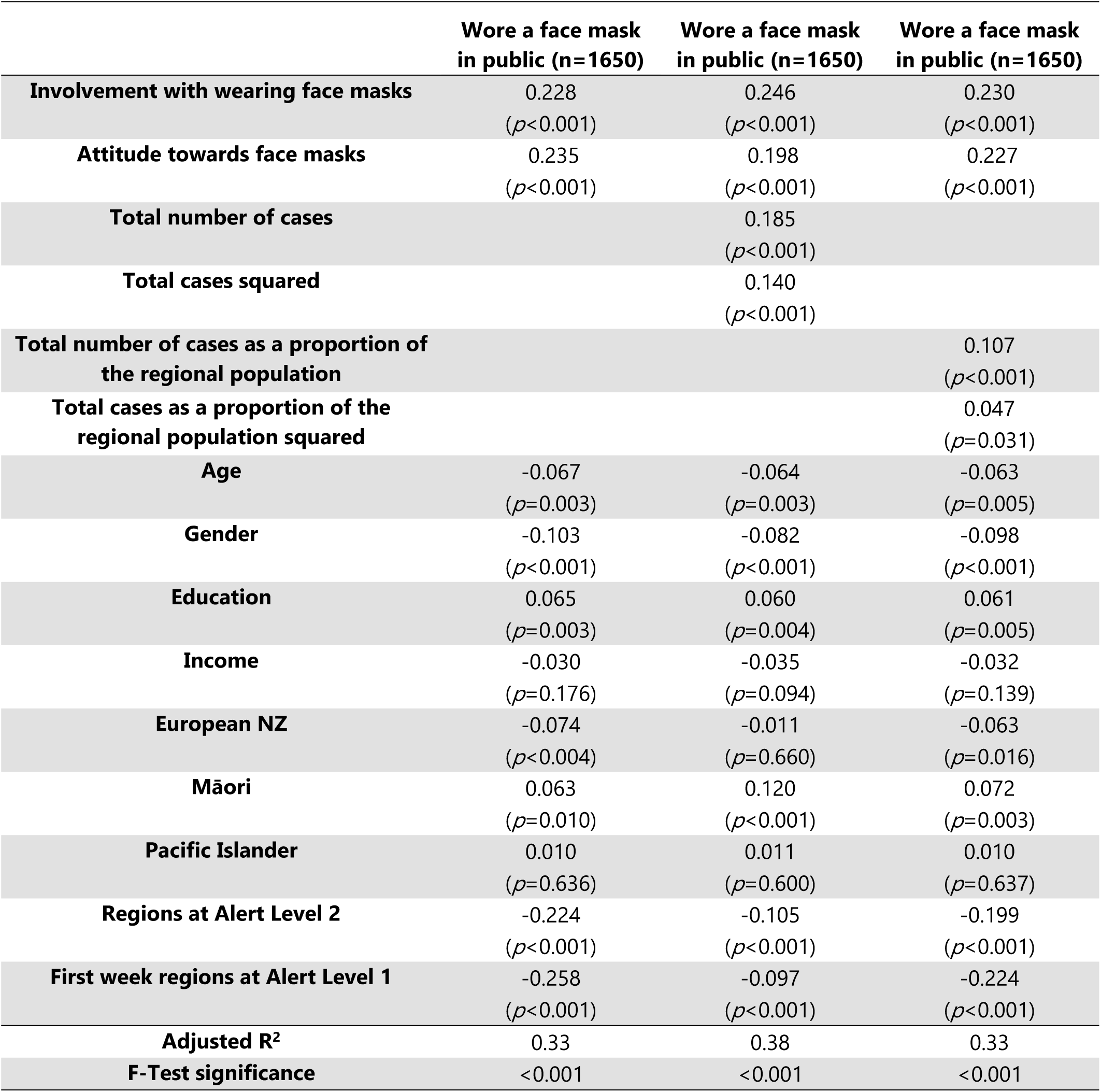
Standardised parameter estimates for wearing face masks in public.

|  | **Wore a face mask**<br>**in public (n=1650)** | **Wore a face mask**<br>**in public (n=1650)** | **Wore a face mask**<br>**in public (n=1650)** |
| --- | --- | --- | --- |
| **Involvement with wearing face masks** | 0.228<br>(*p*<0.001) | 0.246<br>(*p*<0.001) | 0.230<br>(*p*<0.001) |
| **Attitude towards face masks** | 0.235<br>(*p*<0.001) | 0.198<br>(*p*<0.001) | 0.227<br>(*p*<0.001) |
| **Total number of cases** |  | 0.185<br>(*p*<0.001) |  |
| **Total cases squared** |  | 0.140<br>(*p*<0.001) |  |
| **Total number of cases as a proportion of**<br>**the regional population** |  |  | 0.107<br>(*p*<0.001) |
| **Total cases as a proportion of the**<br>**regional population squared** |  |  | 0.047<br>(*p*=0.031) |
| **Age** | -0.067<br>(*p*=0.003) | -0.064<br>(*p*=0.003) | -0.063<br>(*p*=0.005) |
| **Gender** | -0.103<br>(*p*<0.001) | -0.082<br>(*p*<0.001) | -0.098<br>(*p*<0.001) |
| **Education** | 0.065<br>(*p*=0.003) | 0.060<br>(*p*=0.004) | 0.061<br>(*p*=0.005) |
| **Income** | -0.030<br>(*p*=0.176) | -0.035<br>(*p*=0.094) | -0.032<br>(*p*=0.139) |
| **European NZ** | -0.074<br>(*p*<0.004) | -0.011<br>(*p*=0.660) | -0.063<br>(*p*=0.016) |
| **Māori** | 0.063<br>(*p*=0.010) | 0.120<br>(*p*<0.001) | 0.072<br>(*p*=0.003) |
| **Pacific Islander** | 0.010<br>(*p*=0.636) | 0.011<br>(*p*=0.600) | 0.010<br>(*p*=0.637) |
| **Regions at Alert Level 2** | -0.224<br>(*p*<0.001) | -0.105<br>(*p*<0.001) | -0.199<br>(*p*<0.001) |
| **First week regions at Alert Level 1** | -0.258<br>(*p*<0.001) | -0.097<br>(*p*<0.001) | -0.224<br>(*p*<0.001) |
| **Adjusted R<sup>2</sup>** | 0.33 | 0.38 | 0.33 |
| **F-Test significance** | <0.001 | <0.001 | <0.001 |

**Table 12.**
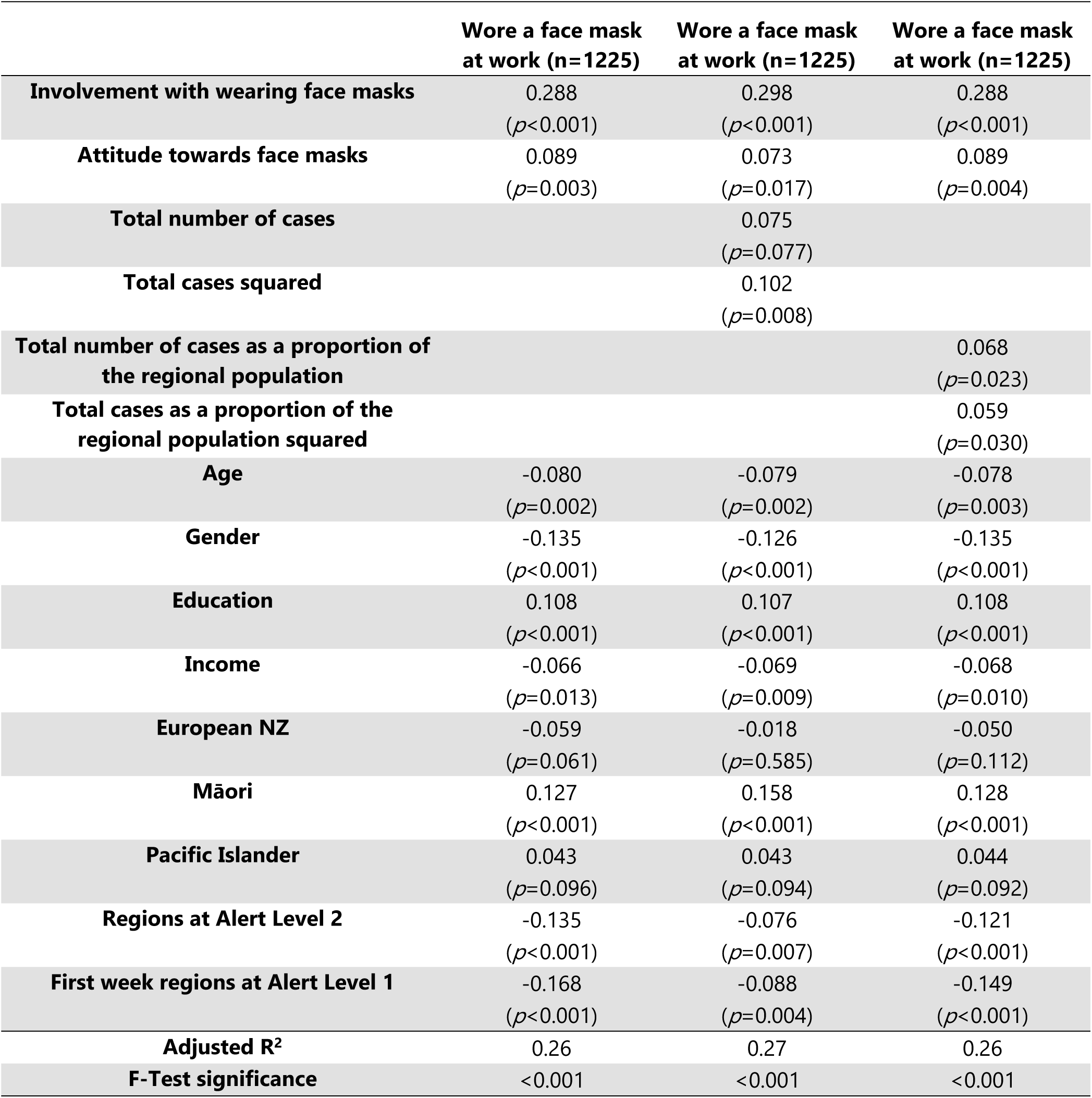
Standardised parameter estimates for wearing face masks at work.

|  | **Wore a face mask**<br>**at work (n=1225)** | **Wore a face mask**<br>**at work (n=1225)** | **Wore a face mask**<br>**at work (n=1225)** |
| --- | --- | --- | --- |
| **Involvement with wearing face masks** | 0.288<br>(*p*<0.001) | 0.298<br>(*p*<0.001) | 0.288<br>(*p*<0.001) |
| **Attitude towards face masks** | 0.089<br>(*p*=0.003) | 0.073<br>(*p*=0.017) | 0.089<br>(*p*=0.004) |
| **Total number of cases** |  | 0.075<br>(*p*=0.077) |  |
| **Total cases squared** |  | 0.102<br>(*p*=0.008) |  |
| **Total number of cases as a proportion of**<br>**the regional population** |  |  | 0.068<br>(*p*=0.023) |
| **Total cases as a proportion of the**<br>**regional population squared** |  |  | 0.059<br>(*p*=0.030) |
| **Age** | -0.080<br>(*p*=0.002) | -0.079<br>(*p*=0.002) | -0.078<br>(*p*=0.003) |
| **Gender** | -0.135<br>(*p*<0.001) | -0.126<br>(*p*<0.001) | -0.135<br>(*p*<0.001) |
| **Education** | 0.108<br>(*p*<0.001) | 0.107<br>(*p*<0.001) | 0.108<br>(*p*<0.001) |
| **Income** | -0.066<br>(*p*=0.013) | -0.069<br>(*p*=0.009) | -0.068<br>(*p*=0.010) |
| **European NZ** | -0.059<br>(*p*=0.061) | -0.018<br>(*p*=0.585) | -0.050<br>(*p*=0.112) |
| **Māori** | 0.127<br>(*p*<0.001) | 0.158<br>(*p*<0.001) | 0.128<br>(*p*<0.001) |
| **Pacific Islander** | 0.043<br>(*p*=0.096) | 0.043<br>(*p*=0.094) | 0.044<br>(*p*=0.092) |
| **Regions at Alert Level 2** | -0.135<br>(*p*<0.001) | -0.076<br>(*p*=0.007) | -0.121<br>(*p*<0.001) |
| **First week regions at Alert Level 1** | -0.168<br>(*p*<0.001) | -0.088<br>(*p*=0.004) | -0.149<br>(*p*<0.001) |
| **Adjusted R<sup>2</sup>** | 0.26 | 0.27 | 0.26 |
| **F-Test significance** | <0.001 | <0.001 | <0.001 |

The regressions where the perception of the risk of infection was assumed to be proportional to the total number of COVID-19 cases were superior to the regressions where the perception of the risk of infection was assumed to be proportional to the total number of COVID-19 cases expressed as a fraction of the population of the region.

Willingness to wear face masks in public was strongly and positively influenced by involvement with, and attitudes towards, wearing face masks. As was the case with behavioural intentions, the variation in respondents’ wearing of face masks in public was only weakly related to their demographic characteristics. Changes in Alert Levels did appear to have some influence on the wearing of face masks in public by regional respondents, though this influence was weakened when the variables intended to account for respondents’ perception of the risk of COVID-19 infection were included in the regression. As hypothesised, the cumulative number of COVID-19 cases in a region had a positive influence on the wearing of face masks in public, and this influence strengthened as case numbers increased.

Willingness to wear face masks at work was positively influenced by involvement with, and attitudes towards, wearing face masks though the effects of attitude are weaker than was the case for wearing face masks in public. The variation in respondents’ wearing of face masks at work was related to gender, education and, slightly, ethnicity. Changes in Alert Levels did appear to influence the wearing of face masks at work by regional respondents. The variables intended to account for respondents’ perception of the risk of COVID-19 infection had a much smaller influence on mask wearing at work compared to their influence on wearing face masks when out in public, perhaps reflecting the lower degree of autonomy in decision-making that individuals have in the workplace.

These results support our hypothesis that the difference between Auckland and regional respondents in the wearing of face masks can be attributed to differences in perceptions of the risk of infection. If regional respondents did perceive the risk of infection from COVID-19 to be lower than Auckland respondents, then they should be less likely than Auckland respondents to seek testing for COVID-19 unless they felt unwell. That is to say, the proportion of respondents who felt unwell when they were tested for COVID-19 should be significantly higher among regional respondents than Auckland respondents. We found this to be the case with approximately 69.7% of regional respondents feeling unwell when they were tested for COVID-19 compared to 50.4% of Auckland respondents (*p*<0.001).

## Discussion

The results are consistent with our hypothesis that the difference between Auckland and regional respondents in their self-reported frequency for wearing face masks when out in public is due to differences in their perceptions of the risk of infection. Both samples of respondents had similar levels of involvement with, and attitudes towards, eliminating COVID-19 from New Zealand. Consequently, both samples displayed similar intentions to:

• take responsibility for eliminating COVID-19 from New Zealand
• change normal behaviour to eliminate COVID-19 from New Zealand
• work with others to eliminate COVID-19 from New Zealand
• make sacrifices to eliminate COVID-19 from New Zealand

Both samples of respondents had similar levels of involvement with, and attitudes towards, self-isolating if unwell or being instructed to do so by a health authority. These similarities resulted in similar intentions to self-isolate if unwell or instructed to do so by a health authority.

Both samples of respondents exhibited similar levels of involvement with, and attitudes towards, wearing face masks when out in public. However, while these similarities might mean both samples had similar intentions to wear face masks, this did not translate into similar behaviour. Regional respondents reported much lower frequency of mask wearing than Auckland respondents. Our results suggest this difference was primarily due to differences in perceptions of the risk of infection as measured by the total number of infections in a region at the time the surveys were conducted (assuming respondents in both samples had similar perceptions of the severity of the health consequences should they contract COVID-19). Differences between the samples with respect to demographic characteristics, and Alert Levels at the time of the surveys, appeared to play a secondary role in explaining the difference between the samples in wearing of face masks.

The willingness of people to adopt behaviours to prevent the spread of COVID-19 such as wearing face masks, self-isolating and getting tested for COVID-19 has been the subject of numerous studies [6,7, 34, 35, 36, 37, 38, 39, 40, 41]. These studies have shown that willingness to adopt these behaviours does depend on people’s attitudes towards them, which in turn depend on their beliefs about the behaviours. Consequently, many of these studies recommend that the adoption of preventative behaviours can be improved through promotional efforts intended to change beliefs and attitudes. Our results have three important implications for such recommendations.

The first concerns the fact that these findings are a reminder that intentions do not always immediately translate into actions. While it is undoubtedly true that changing attitudes can change behaviour, promotional efforts intended to change beliefs and attitudes about preventative behaviours are unlikely to meet with complete success unless health authorities also seek to identify the factors that:

• trigger the translation of intentions into actions, and
• prevent those who are involved to act from acting.

For example, with respect to wearing face masks our results suggest that respondents to our surveys may have relied on the number of COVID-19 infections in their region, together with changes in Alert levels, as signals to trigger the translation of intention into action. This means that providing timely and easily accessible information on the number of infections resulting from community transmission in each region is important. Relatedly, it also means providing timely and easily understandable information on the danger to health posed by different variants of COVID-19 is essential if the public is to make a reasonable judgement about the number of infections from community transmission required to trigger action. This is supposing that there is a relationship, most likely an inverse relationship, between the seriousness of the health risk posed by a variant and the threshold for infections by community transmission below which intentions remain just that, intentions.

Second, bearing in mind the difficulties of engaging with those who are not involved and getting them to observe measures to prevent or slow the spread of COVID-19 [6], our results reinforce the importance of ensuring that measures to prevent the spread of COVID-19 are as simple and convenient to adopt as possible. Improving the ease and convenience of adopting preventative behaviours may, in fact, be more effective in changing behaviour than promotional efforts aimed at changing people’s beliefs and attitudes. In the context of measures to prevent the spread of COVID-19, this translates into ensuring that compliance requires as little effort and thought as practical and is as stress-free as possible [21]; for example, by supplying face masks for free on public transport and in other high-risk locations such as supermarkets [34]. See West et al. [42] for a discussion about the range of interventions that might accompany promotion.

We found, as Gray et al. [43] did, that people’s willingness to wear face masks depended on their beliefs about the effectiveness of face masks in protecting them from infection. Consequently, they recommended developing and promoting to the public clear guidelines on wearing face masks and increasing promotional efforts dispelling myths about the efficacy of masks [43]. While such promotional efforts may meet with success in encouraging intentions, they are only part of the story. Providing timely, trustworthy, and easily accessible information on the number of infections resulting from community transmission in each region is also critical.

Third, it is important to bear in mind that, for New Zealanders at least, wearing a face mask when out in public means constantly disrupting routine behaviours. Consequently, wearing a face mask requires much more time and effort than getting vaccinated for COVID-19, a non-routine action that only needs to be performed two or three times. This suggests that the perceived risk of infection that triggers the translation of the intention to wear a face mask into action is likely to be higher (ceteris paribus) than that required to trigger the translation of the intention to get vaccinated for COVID-19 into action.

Relatedly, this suggests that, once the number of infections by community transmission begin to decline, the public will desire an end to measures to prevent the spread of COVID-19 that constantly disrupt routine behaviours far earlier than for other, less unsettling measures. This means, for instance, that the observance of measures mask wearing, social distancing, using tracer apps and showing vaccine passes is likely to deteriorate even though measures such as being vaccinated and tested are still strongly supported. Ironically, success in vaccinating the public may well encourage the faster abandonment of mask wearing and social distancing if vaccinations are perceived to reduce the risk of transmission and the severity of symptoms.

This raises the possibility that, because misinformation can undermine compliance with COVID-19 measures [44, 45], the distribution of misinformation about the health risks posed by COVID-19 through social media could lead to measures that disrupt daily routines being adopted more slowly, and abandoned more rapidly, than is desirable. This is because such misinformation may provide a self-serving rationale for failing to continue to comply with measures that require a constant investment of time and effort. This reinforces the importance of providing timely, accurate and trustworthy regional information on the spread of COVID-19 variants by community transmission and the severity of the health dangers posed by each variant. These considerations suggest that government authorities must be mindful, when it comes to investing resources in combating misinformation about COVID-19, of the relative importance of combating misinformation targeting people’s beliefs and attitudes and misinformation targeting triggers to action.

Our findings are subject to a several qualifications including the following. First, as the survey samples were drawn from internet-based consumer panels there may be selection bias. While the nature and severity of this bias in relation to the attitudes and involvement we investigated is unknown, it does seem reasonable to suppose, ceteris paribus, that people with low-to-mild involvement may be under-represented in the sample.

Second, as the scales measuring the wearing of face masks, willingness to self-isolate and being tested for COVID-19 were self-reported, our measurements of these behaviours may have been affected by social desirability bias [46]. While Daoust et al. [47] found that social desirability bias appeared to be consistent across gender, age, and education categories, the potential for bias in self-reporting of socially desirable behaviours (or opinions) in the context of the differing degrees of involvement is less clear. However, while there may be a correlation between intensity of involvement and social desirability bias, the dramatic difference between Auckland and regional respondents in their self-reported frequency of wearing face masks suggests that the degree of social desirability bias in our study is small. See Kaine et al. [6] for a more detailed discussion of this matter.

Third, the adoption of behaviours such as the wearing of face masks has been associated with a range of variables including feelings of stress in relation to COVID-19 [21]. We did not include such variables in our analysis, and, while the correlation between these variables and involvement is unknown, it is likely to be positive.

Fourth, the adoption of preventative behaviours such as the wearing of face masks and social distancing has been associated with a range of psychological traits such as pro-sociability and empathy [48, 49, 50]. The correlation between involvement and these psychological traits, and the direction of causation between them, deserves further study.

## Conclusions

In this paper we analysed data from two regional surveys about people’s intentions and behaviour with respect to preventing the spread of COVID-19 in New Zealand. While motivations and intentions were similar across the regions, there was a marked difference in action across the regions, specifically with respect to the frequency of wearing face masks. We found that the translation of intention (preventing the spread of COVID-19) into action (as measured by self-reported frequency of face mask use) was strongly associated with perceptions of the risk of infection (as measured by regional case numbers).

The results serve as a reminder of the importance when designing policies of distinguishing the factors that might influence the formation of behavioural intentions from those that might influence the implementation of those intentions.

## Data Availability

All relevant data are within the manuscript and its Supporting Information files.

## Acknowledgements

We would sincerely like to thank those people throughout New Zealand who completed our questionnaires. Thanks also to our two anonymous referees for their time, patience, constructive advice.

## Supporting information

**S1 File. Questionnaire**

**S2 File. Data**

## Notes

### Competing Interest Statement

The authors have declared no competing interest.

### Funding Statement

This research was funded by the New Zealand Ministry for Business, Innovation and Employment (https://www.mbie.govt.nz/) through the Te Pūnaha Matatini – NZ COVID Modelling Programme (https://www.tepunahamatatini.ac.nz/). MWLR Client project number: UOAX1941. The funders had no role in study design, data collection and analysis, decision to publish, or preparation of the manuscript.

### Author Declarations

The research approach was reviewed and approved by the Manaaki Whenua – Landcare Research’s social ethics process (application no. 2021/27) which is based on the New Zealand Association of Social Science Research code of ethics. Consent was obtained by completion of the questionnaire as participants were randomly recruited from a consumer internet panel. The data were analysed anonymously.

